# A Spatiotemporal Analysis of Socio-Environmental Patterns in Severe Maternal Morbidity: A Retrospective Birth Cohort

**DOI:** 10.1101/2021.03.16.21253540

**Authors:** Stella Harden, Jennifer Runkle, Margaret Sugg

**Affiliations:** Department of Geography & Planning, Appalachian State University, Boone, NC, USA; North Carolina Institute for Climate Studies, North Carolina State University, NC, USA

**Keywords:** Maternal Health, Severe Maternal Morbidity, Cluster Analysis, Social Determinants of Health, GIS

## Abstract

**Objectives:** Severe Maternal Morbidity (SMM) is a group of pregnancy complications in which a woman nearly dies. Despite its increasing prevalence, there is little research that evaluates geographic patterns of SMM and the underlying social determinants that influence excess risk. This study examines the spatial clustering of SMM across South Carolina, US, and its associations with place-based social and environmental factors.

**Methods:** Hospitalized deliveries from 1999 to 2017 were analyzed using Kulldorff’s spatial scan statistic to locate areas with abnormally high rates of SMM. Patients inside and outside risk clusters were compared using Generalized Estimating Equations (GEE) to determine underlying risk factors.

**Results:** Final models revealed that the odds of living in a high-risk cluster were 84% higher among Black patients (OR=1.84, p<.001), 30% higher among Hispanic and Latina patients (OR=1.3, p<.05), and 1.51 times more likely among women living in highly segregated and poorer minority communities (OR=1.51 p<.001). Odds for residing in a high-risk cluster were 23% higher for those who gave birth during a period with temperatures above 30.65°C/87.3°F (OR=1.23, p<.001).

**Conclusions:** This study is the first to characterize the geographic clustering of SMM risk in the US. Our geospatial approach contributes a novel understanding to factors which influence SMM beyond patient-level characteristics and identifies the impact of systemic racism on maternal morbidity. Findings address an important literature gap surrounding place-based risk factors by explaining the contextual social and built environment variables that drive SMM risk.

**Significance:** It is not entirely clear why SMM is increasing in the US. Underlying health conditions, environmental, and social factors have been linked to higher SMM risk. This study is the first to assess these factors across space to determine the characteristics and locations where SMM likelihood is elevated.

**Highlights:**

- First study to examine spatial patterning of severe maternal morbidity (SMM)
- SMM is geographically clustered and increasing in South Carolina, USA
- Highest risk for SMM in racially segregated low-income communities
- Obesity and race were significant individual risk factors
- High ambient temperatures corresponded with high SMM clustering

## Introduction

Severe Maternal Morbidity (SMM) rates rose over 200% in the United States from 1993 to 2014, despite increasing healthcare costs and reductions of SMM in other high-income countries (Centers for Disease Control and Prevention, 2020; American Medical Association, 2020; Geller et al., 2018; Creanga et al., 2014). SMM is a composite group of maternal conditions that signifies when a woman nearly dies from pregnancy complications (Callaghan et al., 2012). Some studies have linked growing rates to individual factors like advanced maternal age, obesity, and comorbidities such as hypertension and diabetes (Grobman et al., 2014). While others have concluded that SMM increase is linked to the increasing prevalence of cesarean delivery (Guglielminotti et al., 2018a; Leonard et al., 2019). Existing studies have shown racial minorities (Admon et al., 2018; Howell et al., 2016), residents in rural communities (Lisonkova et al., 2016; Laditka et al., 2006), and older women (Booker et al., 2018; Leonard et al., 2019) experience elevated SMM rates. Recent research has focused on the role of social and health determinants in influencing SMM (Liese et al., 2019; Guglielminotti et al., 2018b). Yet, it is not entirely clear why SMM is increasing and unevenly impacting certain populations.

There has been limited research on temporal or geographic SMM trends specific to the southeastern US. Recent data show that SMM deliveries were more likely in the South, including South Carolina (SC), and the Northeast (Fingar et al., 2018). Black women in the South have the highest proportion of maternal morbidity in the US (Liese et al., 2019). Individual SMM indicators have been connected to environmental phenomena such as heat waves and seasonal air pollution, but SMM’s environmental impact has not yet been wholly evaluated (Assibey-Mensah et al., 2019; Cil and Cameron, 2017).

While limited research has examined geographic disparities in SMM risks, exploration of spatial patterning of SMM risks could identify hotspots of risk and shed insight on the contextual factors driving these clusters. To our knowledge, no studies have examined the geographic clustering of SMM risk, nor have they identified underlying risk factors that enhance the clustering of SMM. The objective of this study is to identify spatial and temporal SMM trends and relate these patterns to place-based health and environmental determinants that drive high-risk clustering in SC from 1999 to 2017. Findings from this study will provide health professionals with information to enhance targeted maternal health interventions and improve understanding surrounding underlying contextual factors that influence high rates of SMM in a community.

## Methods

### Delivery Discharge Data

We performed a case-only analysis drawing from a retrospective birth cohort study. De-identified patient information was provided by the SC Department of Health and Environmental Control from 1999 to, 2017 (*n*=860,535). SMM was categorized using the CDC definition which includes twenty-one procedures and diagnoses adapted from the ninth revision of the International Classification of Diseases (ICD) codes. A published algorithm was used to flag hospital deliveries as SMM-related hospitalizations and takes into account a number of factors like maternal mortality, delivery mode, and length of stay (CDC, 2020).

### SMM Measures

Previous research has shown that blood transfusion is the most commonly occurring indicator of SMM (Callaghan et al., 2012; Creanga et al., 2014). However, assigned ICD-9-CM procedure codes do not include the number of units of blood transferred and may result in artificially high SMM rates. To address this limitation, the primary study outcomes of interest were operationalized as: 1) SMM composite measure of twenty-one CDC recognized indicators with blood transfusion (SMM21) and 2) SMM without blood transfusion (SMM20). SMM cases were geocoded to their residential zip code based on hospital administration records. Patients with a SMM diagnosis included in the study were between the ages of 10 and 55 years with a gestation of 20 to 44 weeks.

### Patient-Level Factors

The following patient-level factors were provided directly from hospitalized medical records: year of hospital delivery discharge, age, race and ethnicity, insurance type, and zip code. The following conditions were identified using ICD-9-CM diagnosis and procedure codes: prior cesarean, cesarean procedure, delivery type, rupture of uterus, preeclampsia, obesity, hypertension, hysterectomy, eclampsia, diabetes, smoking, depression, bipolar disorders, and asthma. Race and ethnicity was categorized into white, Hispanic or Latino, Black, or other with white patients serving as the referent group. Patient age was sorted into four groups: 19 years and younger 20 to 29 years (referent), 30 to 39 years, and 40 years and older. Insurance type was classified as private, Medicare/Medicaid, self pay, or other with privately insured patients serving as the reference.

### Community-Level Factors

The following data obtained at the zip code-level was used to represent social and environmental factors that may influence SMM likelihood: toxic release inventory (TRI) site emissions, number of primary care facilities, racial residential segregation, income inequality, and racialized economic residential segregation derived from the Index of Concentration at the Extremes (ICE). Temperature maximum and minimum values were collected at the county-level for every year and stratified into quantiles with the lowest group set as the reference. Primary care facility data was provided by the US Health Resources and Services Administration and separated into three categorical groups: zero facilities, one facility, and two or more facilities.

### Deprivation Index

Vulnerable areas can be determined by a deprivation index that measures the impact of human and physical environments across spatial contexts. The Index of Concentration at the Extremes (ICE) measures racial residential segregation, income inequality, and racialized economic residential segregation and was used in this study (Massey, 1996). A value of −1 indicates 100% of the population within a unit is the most deprived while a value of 1 indicates 100% of the population is the most privileged. ICE metrics are generated using three measures: income, race, and a combination of race and income. We hypothesize that trends in SMM will follow patterns of racialized economic segregation since poverty has been linked to decreased maternal care access and comorbidities such as obesity and hypertension (Nagahawatte and Goldenberg, 2008). ICE was calculated at the zip code level using the 2017 American Community Survey (ACS) 5-Year Estimates and 2010 Census data (*Supplementary Table 1*).

### Cluster Analysis

Spatiotemporal clustering methods have yet to be applied to SMM in the US. SaTScan, a descriptive visualization mapping tool, was used to examine the spatial and temporal clustering of SMM21 and SMM20. SaTScan uses Kulldorff’s spatial scan statistic to identify geographic hotspots of elevated risk at user-specified window sizes (Kulldorff et al., 1997). A discrete Poisson model was used to locate high-risk SMM clusters with (SMM21) and without blood transfusions (SMM20). This model was used due to the rareness of SMM relative to all deliveries and because it produces more conservative *p*-values than the alternative Bernoulli model (Root et al., 2009). Clustering analysis was used to detect purely temporal, purely spatial, and space-time clusters using a maximum circular window size set to, 20% of the total at-risk population. A window size of 20% scans for localized clusters relative to the default of 50% and is better for informing targeted interventions.

### GEE Modeling

To determine the impact of individual and community-level factors on SMM risk and to differentiate characteristics between high-risk cluster and non-cluster deliveries, generalized estimating equations (GEE) with logistic regression was used. The models accounted for the association between social and environmental factors and SMM inside and outside of high-risk clusters. The GEE method is best used in longitudinal, nested, or repeated measures study designs (Ballinger, 2004), and has been applied to other maternal health SatScan research (Chong et al., 2013).

GEE with an exchangeable correlation matrix were used to account for the clustering of SMM cases within zip codes. Covariates included in the first model were: race, age, comorbidities, and insurance status. Environmental variables such as TRI emissions, temperature maximums and minimums, ICE measures of racialized and economic segregation, and number of primary care facilities were measured in the second model. The final model incorporated significant factors from both models to determine the most influential characteristics for residing in a high-risk SMM cluster. Using stepwise regression, significant covariates for SMM20 and SMM21 were identified at the 95% confidence level.

## Results

### Patient Characteristics

Between January 1, 1999, and December 31, 2017, there were 8,255 deliveries that resulted in SMM20 and 15,529 characterized by SMM21. These constituted 0.97% and 1.8% of all hospitalized deliveries. The majority of SMM patients were Black despite white patients constituting the population majority (*Table 1*). Black patients accounted for 6,950 (44.8%) of all SMM cases. Blood transfusion, the most frequent indicator, occurred in 56% of SMM deliveries and disproportionately impacted Black and Hispanic women.

**Table 1.** Characteristics for Patients with SMM, South Carolina 1999-2017.

| **Characteristic** | **SMM21**<br>**(n=15529)** | **SMM20**<br>**(n=8251)** | **All Deliveries**<br>**(n=860535)** |
| --- | --- | --- | --- |
| *Race/Ethnicity* |  |  |  |
| White | 6690 (43.1) | 3751 (45.5) | 482707 (56.1) |
| Black | 6952 (44.8) | 3696 (44.8) | 283335 (32.9) |
| Hispanic/Latino | 892 (5.7) | 365 (4.4) | 42416 ( 4.9) |
| Other Race | 995 (6.4) | 439 (5.3) | 52077 ( 6.1) |
| *Age* |  |  |  |
| 19 and Younger | 1900 (12.2) | 836 (10.1) | 101017 (11.7) |
| 20 -, 29 | 7972 (51.3) | 4020 (48.7) | 483006 (56.1) |
| 30 - 39 | 5135 (33.1) | 3049 (37.0) | 260377 (30.3) |
| 40 and Older | 522 (3.4) | 346 (4.2) | 16135 ( 1.9) |
| *Insurance* |  |  |  |
| Medicare/Medicaid | 8659 (55.8) | 4343 (52.6) | 424201 (49.3) |
| Private | 5751 (37.0) | 3334 (40.4) | 369882 (43.0) |
| Self Pay | 488 (3.1) | 229 (2.8) | 25770 ( 3.0) |
| Other | 631 (4.1) | 345 (4.2) | 40682 ( 4.7) |

### Geographic Clustering of SMM Risk

Six significant (*p*< .01) high-risk spatial clusters were present throughout the entire study period for SMM21 (*Figure 1*). Two significant (*p*< .01) high-risk spatial clusters were present throughout the entire study period for SMM20. Purely temporal clusters tracked the incidence of SMM from 1999 to 2017 for the entire study area (*Figure 2)*. Using both a four year and two year maximum cluster size, SMM20 clusters occurred during, 2016 and 2017 (RR=1.78, *p*<.001). SMM21 clusters with a two-year maximum temporal window were identified during, 2016 and 2017 (RR=1.35, *p*<.001), while clusters with a four year maximum window occurred in 2017 (RR=1.33, *p*<.001).

**Figure 1.**
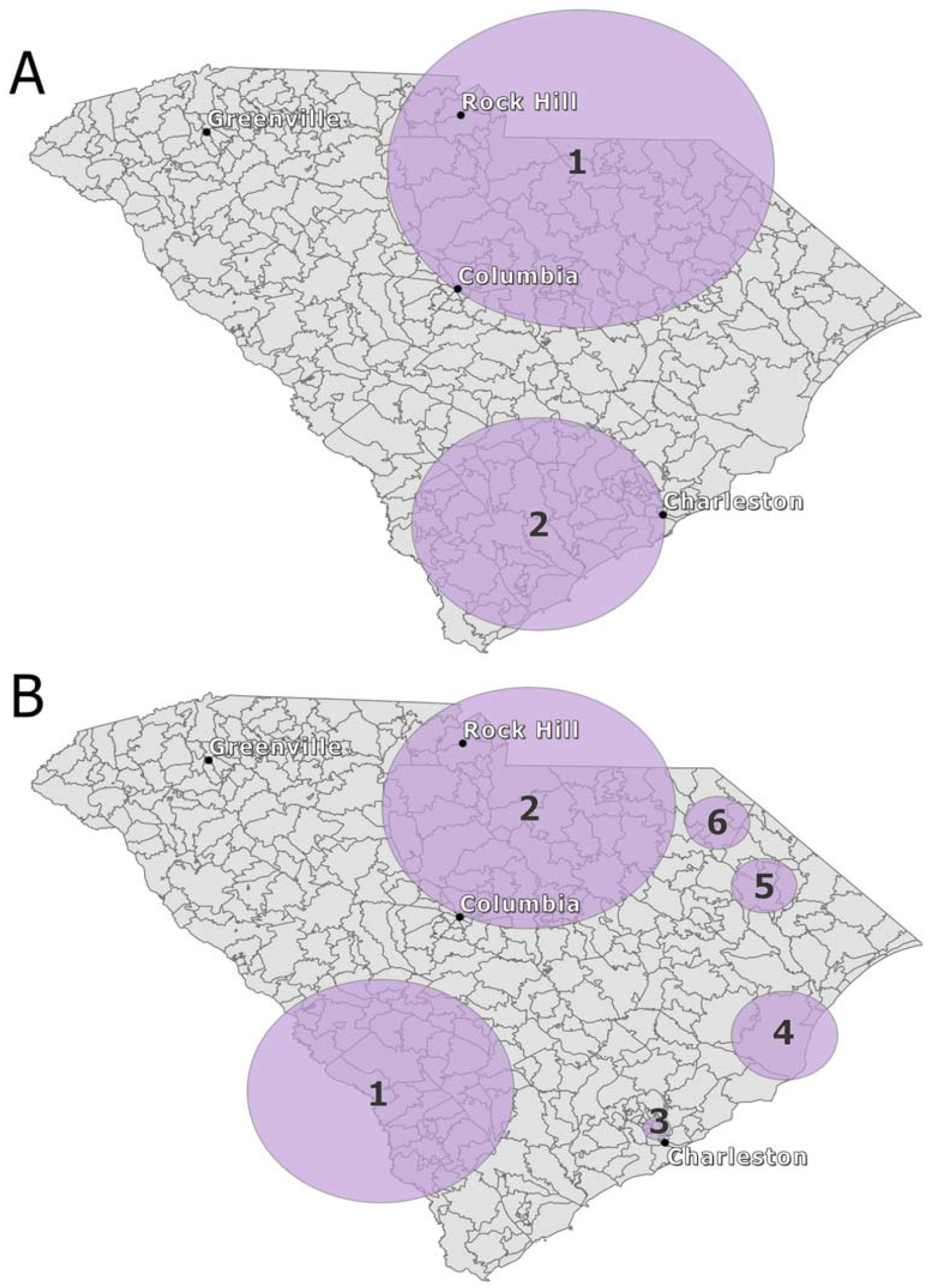
Purely Spatial High-Risk Clusters for, 20% of the Population at Risk. (A) SMM20 (B) SMM21

**Figure 2.**
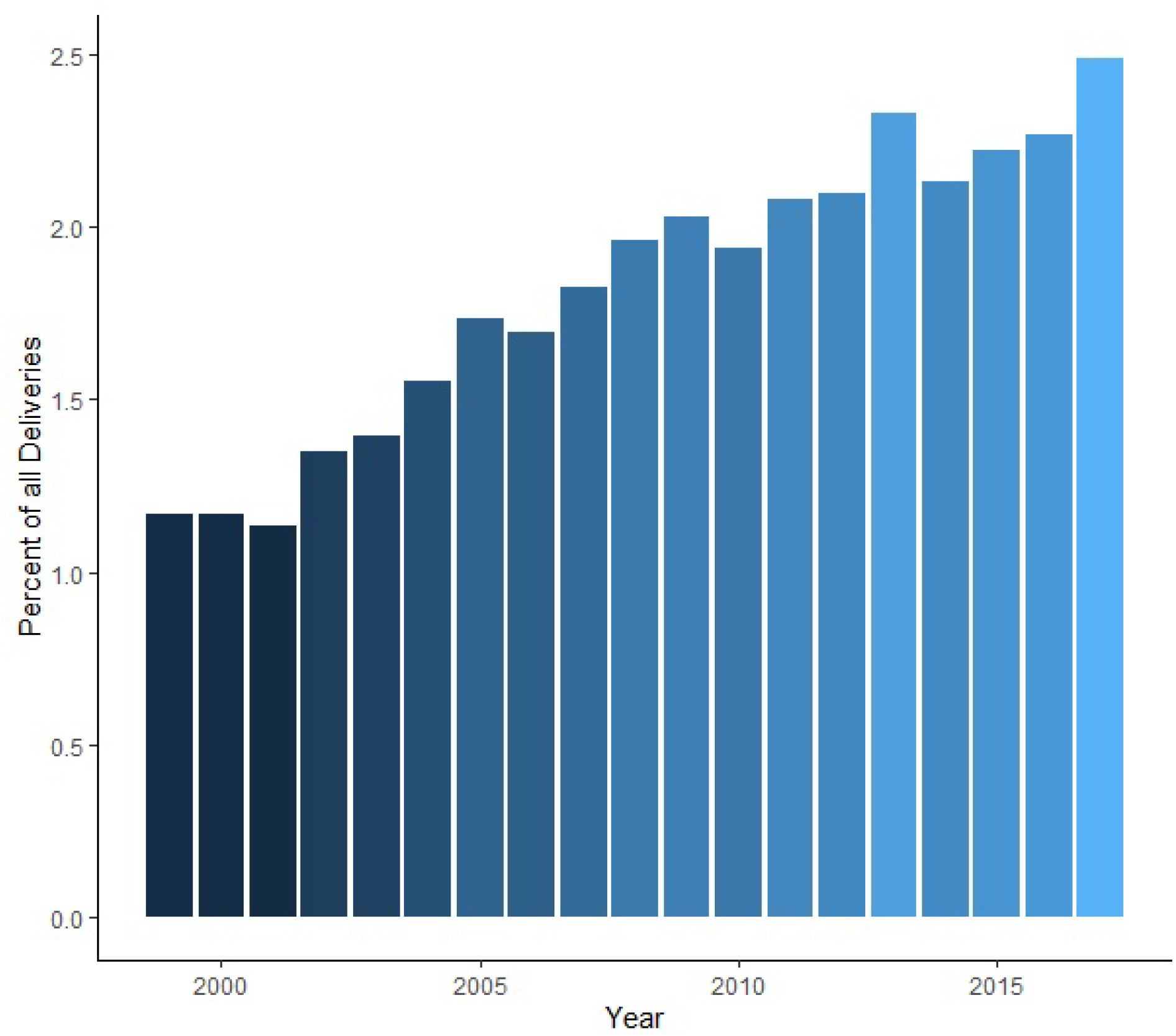
Annual Trends of Hospitalized Deliveries Characterized by SMM (Percentage), South Carolina 1999-2017

A space-time SaTScan analysis using a 20% spatial window scanned for significant clusters (*p*<.01) using 2 year intervals (*Figure 3*). Three space-time clusters for SMM20 were present from 2016 to 2017 in the north and northwest. During the same period, the primary cluster was identified in the southeast (RR=2.19). Three space-time SMM21 clusters were present in different areas of SC than SMM20. The primary SMM21 cluster was found in the south-central region between, 2016 and, 2017 (RR=1.8). Secondary clusters were identified in the north during 2013 and 2014 and in a localized southeastern area between 2007 and 2008.

**Figure 3.**
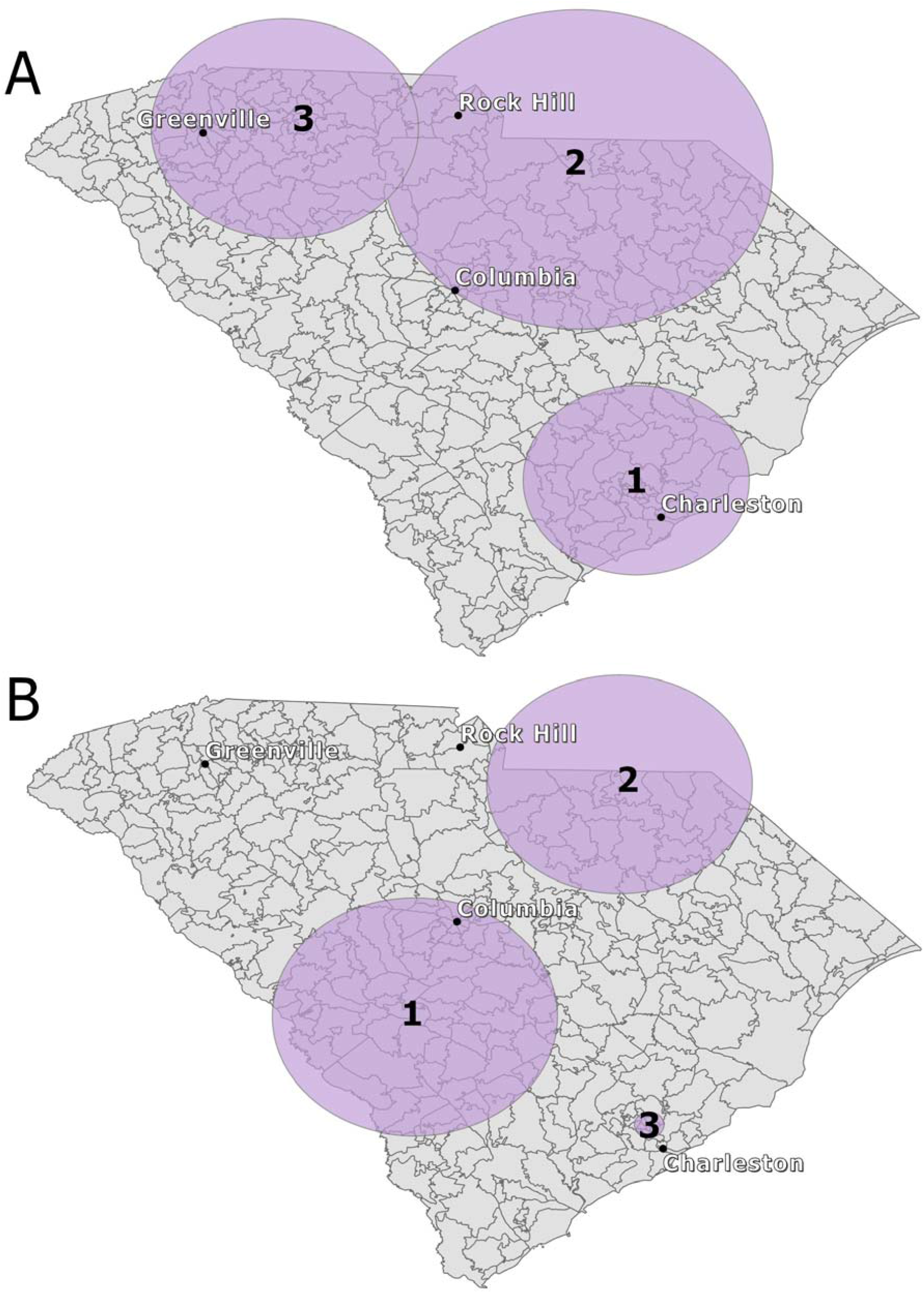
Spacetime High-Risk Clusters for, 20% of the Population at Risk. (A) SMM20 (B) SMM21

Patient and place-based risk factors varied for women with SMM who resided inside clusters and outside of clusters. Results showed that 62.5% of high-risk SMM21 clusters were composed of Black patients. Hispanic and Latino women were more likely to reside in an SMM20 cluster. A total of 60.6% of women with a SMM21 event lived in a zip code characterized by extreme deprivation and 42.2% of women with a SMM20 event lived in the most deprived zip codes (*Figure 4*).

**Figure 4.**
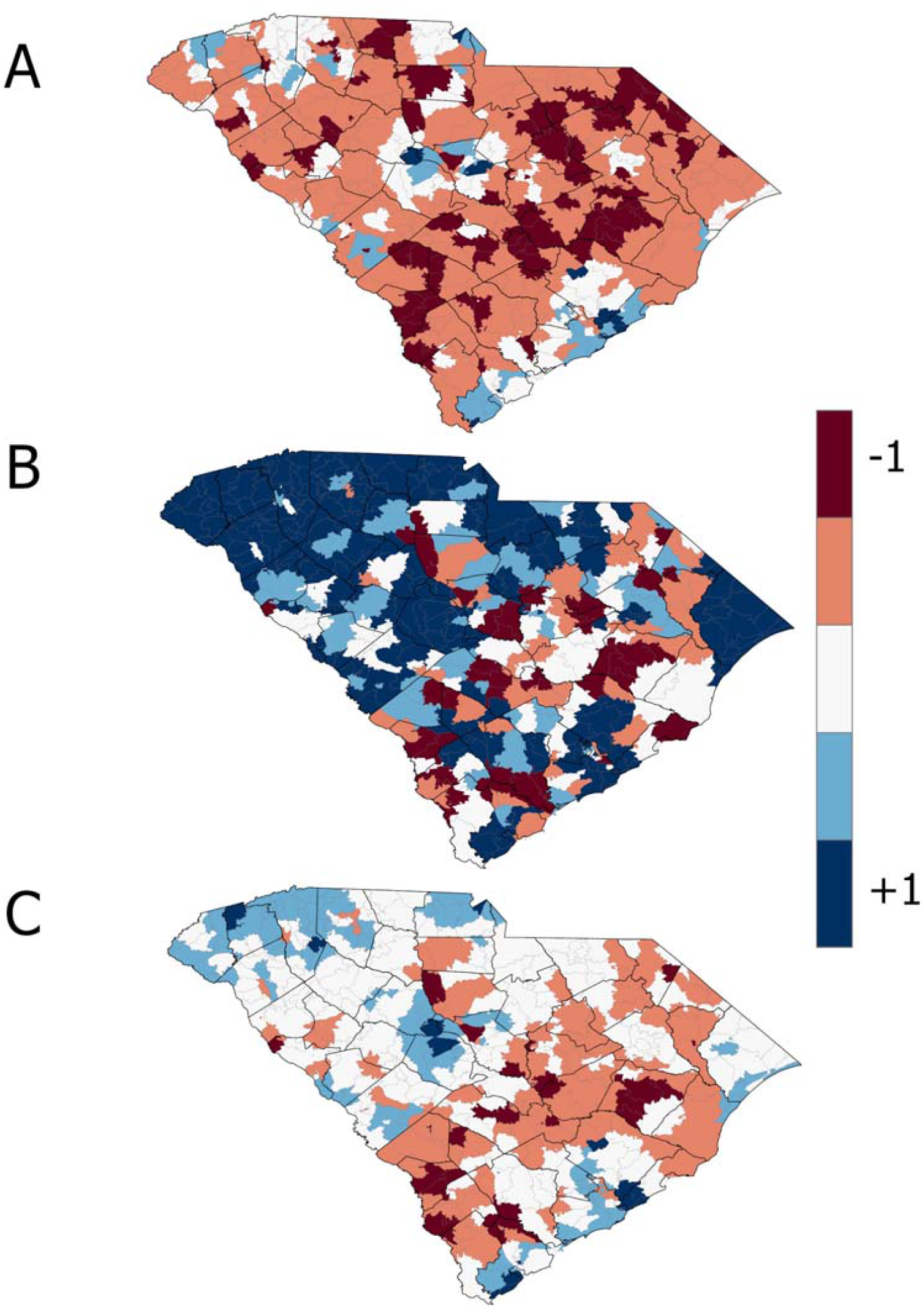
ICE Metrics for 2017. −1 represents everyone in the most deprived group and +1 represents everyone in the most privileged group. (A) Income (B) Race (C) Income and Race. The ICE metric highlighted locations of extreme racial and economic residential segregation in the more privileged western region and more deprived southeast with the exception of urban centers.

### SMM20 Risk Factors

GEE models determined significant covariates for SMM20 cluster risk (*Table 2*). The odds of living in a high-risk SMM cluster was significantly higher for Black women (OR=2.3, *p*<.001), Hispanic or Latina women (OR=1.43, *p*<.01), and those presented with obesity (OR=1.28, *p*<.001). Upon relating community-level factors to SMM clustering, we observed that the odds of living in a high-risk cluster were 45% higher for women who gave birth during temperature minimums of at least 19.27°C/66.7°F (OR=1.45, *p*<.01) and 7.85 times higher for those living in the most racially segregated zip codes (OR=7.85, *p*<.001). Final models combining patient and community factors revealed that the odds of living in a high-risk cluster was 84% higher among Black patients (OR=1.84, *p*<.001), 30% higher among Hispanic and Latina patients (OR=1.3, *p*<.05), and 1.51 times more likely among women living in highly segregated and poorer minority communities (OR=1.51 *p*<.001) compared to those in low-risk clusters. Odds for residing in a high-risk cluster were 31% higher for women with a pre-existing diagnosis of obesity (OR=1.31, *p*<.001) and, 23% higher for those who gave birth during a period with temperature maximums above 30.65°C/87.3°F (OR=1.23, *p*<.001) compared to women in low-risk clusters.

**Table 2.** SMM20 GEE Models, * *p*-value <.05 ** p-value <.01 *** p-value <.001

| Model 1: |  |  | Model, 2: |  |  | Model 3: |  |  |
| --- | --- | --- | --- | --- | --- | --- | --- | --- |
| Predictors | OR | CI | Predictors | OR | CI | Predictors | OR | CI |
| Obesity*** | 1.28 | 1.11–1.47 | Temperature<br>Minimum Group, 2* | 1.19 | 1.00–1.41 | Obesity*** | 1.31 | 1.13 – 1.52 |
| *Race/Ethnicity* |  |  | Temperature<br>Minimum Group 3 | 1.11 | 0.90 – 1.38 | Black*** | 1.84 | 1.65 –, 2.05 |
| Black*** | 2.3 | 2.0 –2.54 | Temperature<br>Minimum Group 4** | 1.45 | 1.14 – 1.84 | Hispanic/<br>Latino* | 1.3 | 1.03 – 1.64 |
| Hispanic/<br>Latino** | 1.43 | 1.14–1.79 | Temperature<br>Maximum Group, 2 | 1.07 | 0.91 – 1.27 | Medicare/<br>Medicaid* | 0.89 | 0.81 – 0.99 |
| White | 1.0 | (ref) | Temperature<br>Maximum Group 3 | 1 | 0.81 – 1.23 | Self Pay* | 0.76 | 0.58 – 0.98 |
| *Insurance* |  |  | Temperature<br>Maximum Group 4 | 0.89 | 0.70 – 1.14 | Temperature<br>Maximum<br>Group, 2* | 1.14 | 1.01 – 1.29 |
| Medicaid/<br>Medicare | 0.96 | 0.87–1.05 | No Primary Care<br>Facilities*** | 0.03 | 0.02 – 0.07 | Temperature<br>Maximum<br>Group 3* | 1.14 | 1.01 – 1.28 |
| Self Pay* | 0.76 | 0.59–0.98 | One Primary Care<br>Facility*** | 0.07 | 0.03 – 0.15 | Temperature<br>Maximum<br>Group 4*** | 1.23 | 1.09 – 1.39 |
| Privately | 1.0 | (ref) | Race ICE Group<br>1*** | 7.85 | 6.78 – 9.09 | No Primary<br>Care<br>Facilities*** | 0.04 | 0.02 – 0.08 |
|  |  |  | Race ICE Group,<br>2*** | 4.25 | 3.74 – 4.82 | One Primary<br>Care<br>Facility*** | 0.08 | 0.04 – 0.18 |
|  |  |  | Income ICE Group<br>1*** | 0.28 | 0.24 – 0.32 | Race and<br>Income ICE<br>(Most<br>Deprived)*** | 1.51 | 1.33 – 1.71 |
|  |  |  | Income ICE Group,<br>2*** | 0.46 | 0.40 – 0.52 | Race and<br>Income ICE<br>(Middle<br>Group)* | 0.86 | 0.76 – 0.98 |

### SMM21 Risk Factors

The odds of living in a high-risk SMM cluster was significantly higher for Black women (OR=2.6, *p*<.001) and those enrolled in Medicaid or Medicare (OR=1.13, *p*<.01) compared to those outside of clusters (*Table 3*). Upon relating community-level factors to SMM clustering, we found that the odds of living in a high-risk cluster were 8.6 times higher for women in the most racial segregated zip codes (OR=8.56, *p*<.001) and, 20% higher for those living in more extreme poverty areas (OR=1.2, *p*<.01) compared to those outside high-risk SMM clusters. The final model combining patient and community factors revealed that the odds of residence in a high-risk cluster was 6.7 times higher for women living in highly segregated, poor areas (OR=6.69, *p*<.001) and 43% higher among Black patients (OR=1.43 *p*<.001).

**Table 3.** SMM21 GEE models, * *p*-value <.05 ** p-value <.01 *** p-value <.001

| Model 1: |  |  | Model, 2: |  |  | Model 3: |  |  |
| --- | --- | --- | --- | --- | --- | --- | --- | --- |
| Predictors | OR | CI | Predictors | OR | CI | Predictors | OR | CI |
| Black*** | 2.6 | 2.40 –, 2.83 | No Primary Care Facilities*** | 0.08 | 0.06 – 0.12 | Black*** | 1.43 | 1.30 – 1.57 |
| Hispanic/Latino | 0.92 | 0.77 –, 1.11 | One Primary Care Facility*** | 0.26 | 0.18 – 0.38 | Hispanic/Latino*** | 0.66 | 0.54 – 0.81 |
| Medicaid/Medicare** | 1.13 | 1.04 –, 1.23 | ICE Race Group 1*** | 8.56 | 7.47 – 9.81 | Medicaid/Medicare | 0.96 | 0.88 – 1.05 |
| Self Pay*** | 0.69 | 0.55 –, 0.87 | ICE Race Group, 2*** | 2.65 | 2.34 – 3.01 | Self Pay*** | 0.6 | 0.46 – 0.77 |
| Smoking* | 0.82 | 0.68 –, 0.99 | ICE Income Group 1** | 1.2 | 1.06 – 1.37 | No Primary Care Facilities*** | 0.07 | 0.05 – 0.11 |
| Hysterectomy* | 0.82 | 0.69 –, 0.96 | ICE Income Group, 2*** | 1.68 | 1.49 – 1.89 | One Primary Care Facility*** | 0.29 | 0.20 – 0.41 |
| Under 19 years | 1.07 | 0.96 –, 1.19 |  |  |  | Race and Income ICE (Most Deprived)*** | 6.69 | 5.88 – 7.61 |
| Between 30 and 39 years* | 0.9 | 0.83 –, 0.98 |  |  |  | Race and Income ICE (Middle Group)*** | 2.33 | 2.05 –, 2.64 |
| Over 40 years | 1.09 | 0.89 –, 1.32 |  |  |  |  |  |  |

## Discussion

This study is the first to characterize the geographic clustering of SMM risk in the US, particularly in a Southern state. Our geospatial approach contributes a novel understanding to factors which influence SMM beyond patient-level characteristics and identifies the impact of systemic racism on maternal morbidity. Findings address an important gap in the maternal morbidity literature surrounding place-based risk factors by explaining the contextual social and built environment variables that drive SMM risk and vary greatly across the state. Results showed significant spatial clustering of SMM risk in geographic regions. Six spatial clusters for SMM21 and two spatial clusters for SMM20 were identified from 1999 to 2017. With the exception of a localized cluster in the southeast in 2008, significant space-time and purely temporal clusters occurred in the final two or four years of the study period. These findings support a growing body of evidence that SMM risk is steadily increasing in the US (Leonard et al., 2019; Fingar et al., 2018; Creanga et al., 2014), and our findings suggest that SMM risks are place-specific.

High-risk SMM clusters were composed of a greater proportion of racial minorities and those in segregated low-income areas than regions outside of clusters. These findings within the study population support existing literature that race and ethnicity are closely linked to SMM risk (Admon et al., 2018; Aziz et al., 2019; Howell et al., 2016). This evidence also suggests that racialized economic segregation strongly influences SMM risk within poorer minority communities. Prior studies have linked systemic discrimination to adverse maternal health and childbirth outcomes (Chambers et al., 2019; Owens and Fett, 2019). Women residing in high-risk spatial SMM clusters were significantly more likely to live in disadvantaged racialized economic areas. Similar associations between higher SMM risk and being non-White and residing in a high poverty area were observed in a previous study (Guglielminotti et al., 2018a). The primary cluster for SMM21 overlapped with more racialized economically disadvantaged areas in the southeast. The northeast and northwest, areas with relatively high or average ICE values for race and income, did not have any SMM high-risk clusters present.

Obesity, a known contributor to adverse maternal health outcomes, was found to only influence SMM excluding blood transfusions in this population. This partially explained the increased likelihood for low income women to have SMM as an overwhelming amount of literature has tied low socioeconomic status with higher rates of obesity (Langenberg et al., 2003). Both high and low body mass index (BMI) slightly increased SMM in a study of Washington state but obesity was not associated with increased risk in a California study (Lisonkova et al., 2017; Leonard et al., 2019). Despite having areas with higher income inequality, it is possible that no spatial SMM clusters were identified in the north due to the success of the Centering Pregnancy program which originated in the area. Centering Pregnancy is a group-based prenatal care model that has been connected to a number of improvements in prenatal care and outcomes, particularly in public health clinics serving low-income populations (Klima et al., 2009). In addition to providing support and pregnancy education, the program has reduced preterm birth by 47% and encouraged public health agencies to invest in greater preventive measures affordable to all patients (Prisma Health).

Though a relationship was not found between TRI emissions and SMM cluster risk in our analysis, it is known that underprivileged populations tend to reside in more hazardous environmental areas compared to more affluent populations (Mohai et al., 2009). Considering this elevated risk, a focus on areas with more racialized economic segregation may identify these place-specific toxic environmental hazards.

High temperatures were associated with SMM excluding blood transfusions. Our conclusions align with those from Cil and Cameron showing that heatwaves negatively impact maternal health (2017). It is likely that a strong correlation exists between SMM and climate extremes as literature has identified a relationship between higher or colder than average seasonal temperatures and adverse birth outcomes (Basu et al., 2010). More research is needed to understand the impacts of climate on SMM.

### Strengths and Limitations

This study examined a large dataset of maternal characteristics for an entire state from 1999 to 2017. We were able to link multiple births to a single mother, which made for a more accurate longitudinal investigation. The inclusion of geographical analysis at a relatively fine scale provided important information on factors that can be leveraged in focused community interventions (Huang et al., 2009). A further strength of our geographical analysis was the inclusion of time by examining space-time scan statistics, which showed spatial clusters occurring in the most recent study years.

There were several limitations in this study. While a useful visualization tool, SaTScan cannot explain the reason for risk variation in clusters. The use of a circular scan window identifies populations that are in circularly shaped areas, however this is not necessarily the pattern that SMM likelihood follows and use of an elliptical window might provide slightly increased capability for scanning along some geographic features (Chong et al., 2013). The 20% spatial window utilized for the GEE models presented risk clusters at a relatively localized scale compared to the 50% default which is more beneficial for intervention in homogenous communities rather than regional initiatives. While an association is likely present between SMM and ambient air pollution linked to seasonality, our analysis was limited by poor air quality data.

## Conclusion

Our initial findings highlight the need for more place-based research to inform the location of interventions at local scales for vulnerable populations within high-risk SMM clusters. Findings revealed that SMM risks increased over time and disproportionately impacted women of color. Results showed significant geographic disparities in SMM risk which was largely driven by economic racialized residential segregation, a measure of structural racism, and obesity.

## Ethical Considerations

Data in this manuscript were based on de-identified hospitalized delivery discharges authorized and ethically provided by the South Carolina Department of Health and Environmental Control. The study was reviewed and approved by an Institutional Review Board.

## Data Availability

These data were obtained through a data use agreement with a state agency data warehouse and are not publicly available.

## Appendix

**Supplemental Table 1.** ICE formulas using Census variables by Krieger et al., 2016

|  |  |
| --- | --- |
| **Income** | (number of persons in high-income households) - (number of persons in low-income households) / total population with household income data |
| **Race** | number of white non-Hispanic persons) - (number of Black non-Hispanic persons) / number of persons with race/ethnicity data |
| **Income and Race** | (number of white non-Hispanic high-income persons) - (number of Black alone low-income persons) / number of persons with race/ethnicity and household income data |

## Notes

### Competing Interest Statement

The authors have declared no competing interest.

### Funding Statement

No external funding was obtained to do this research.

### Author Declarations

The Appalachian State University Institutional Review Board reviewed the study protocol and ruled that the study was category 4 exempt (protocol #: 19-0272).

